# Local Neuronal Sleep after Stroke: The role of cortical bistability in brain reorganization

**DOI:** 10.1101/2024.04.06.24304834

**Authors:** Caroline Tscherpel, Maike Mustin, Marcello Massimini, Theresa Paul, Ulf Ziemann, Gereon R. Fink, Christian Grefkes

## Abstract

**Background:** Acute cerebral ischemia triggers a number of cellular mechanisms not only leading to excitotoxic cell death but also to enhanced neuroplasticity, facilitating neuronal reorganization and functional recovery.

**Objective:** Transferring these cellular mechanisms to neurophysiological correlates adaptable to patients is crucial to promote recovery post-stroke. The combination of TMS and EEG constitutes a promising readout of neuronal network activity in stroke patients.

**Methods:** We used the combination of TMS and EEG to investigate the development of local signal processing and global network alterations in 40 stroke patients with motor deficits alongside neural reorganization from the acute to the chronic phase.

**Results:** We show that the TMS-EEG response reflects information about reorganization and signal alterations associated with persistent motor deficits throughout the entire post-stroke period. In the early post-stroke phase and in a subgroup of patients with severe motor deficits, TMS applied to the lesioned motor cortex evoked a sleep-like slow wave response associated with a cortical off-period, a manifestation of cortical bistability, as well as a rapid disruption of the TMS-induced formation of causal network effects. Mechanistically, these phenomena were linked to lesions affecting ascending activating brainstem fibers. Of note, slow waves invariably vanished in the chronic phase, but were highly indicative of a poor functional outcome.

**Conclusion:** In summary, we found evidence that transient effects of sleep-like slow waves and cortical bistability within ipsilesional M1 resulting in excessive inhibition may interfere with functional reorganization, leading to a less favorable functional outcome post-stroke, pointing to a new therapeutic target to improve recovery of function.

## INTRODUCTION

After stroke, a cascade of repair- and growth-related processes including synaptogenesis, neurogenesis, and axonal sprouting have been identified to occur not only locally in peri-infarct areas but also extend to contralesional regions [1,2]. Crucially, invasive recordings of cortical activity have associated axonal sprouting after ischemia with synchronous low-frequency oscillations, reminiscent of the phenomenon of slow waves during NREM sleep [2–4]. While, at first glance, stroke and sleep may not be unambiguously reconciled, the notion of slow-wave sleep merits further consideration in the context of stroke. Especially, since slow waves have not only been described ubiquitously during sleep but also as a locally confined phenomenon during wakefulness [5,6]. Moreover, both conditions represent a particular environment for neuroplasticity and reorganization with potentially analogous neuronal underpinnings.

In NREM sleep, studies have associated slow waves with neuronal off-periods [7], aggregating the phenomenon of neurons to fall into a hyperpolarized down-state, i.e., a period of suppressed firing after an initial increase in activity, often referred to as cortical bistability [8–10]. Extensively studied in in-vitro, in-vivo, and in-silico models, it has been suggested that this phenomenon is attributable to adaption mechanisms such as activity-dependent potassium currents and increased inhibition [11–13]. In parallel, similar effects, i.e., hyperpolarization, a significant involvement of K-ATP channels, and increased inhibition have all also been shown post-stroke [14,15]

While direct invasive recordings of slow waves remain mostly limited to animal models, the methodology of combined transcranial magnetic stimulation (TMS) and EEG [16] seems to be optimally suited to capture sleep-like slow waves and associated off-periods in humans non-invasively, as the signal arises from the perturbation of intrinsic neuronal activity [9,17].

Crucially, it has also been shown that TMS-EEG can evoke slow wave responses irrespective of the presence of spontaneous slow waves in the background resting EEG [9,18]. Sleep-like slow waves and a breakdown of complex neuronal interactions have been demonstrated in unresponsive wakefulness syndrome, first contextualizing the principles of cortical bistability in the framework of brain lesion [9,19]

For stroke, it is well known that the structural damage profoundly impacts interregional connectivity and the functional network architecture [20]. Yet crucially, TMS-EEG has recently revealed the occurrence of local slow waves in conscious awake stroke patients [18,21]. Furthermore, TMS-EEG responses have been ascribed to inform about the functional deficit and the potential for recovery [21]. While slow-wave sleep has been attributed to neuronal recovery and plasticity [22], and slow waves in stroke were hypothesized to guide axonal sprouting, thereby facilitating reorganization [4,23], bistability and slow waves occurring during wakefulness seem to be functionally detrimental [6,9].

Establishing a systematic link between slow waves and cortical off-periods post-stroke with their behavioral consequences requires longitudinal measurements from the acute to the chronic phase. Therefore, we assessed TMS-EEG early after stroke and followed the course of functional recovery and development of alterations of TMS-EEG responses alongside reorganization to the chronic phase. We hypothesized that TMS-EEG responses disclose behaviorally relevant information about neural reorganization throughout the entire post-stroke period. We specifically tested the hypothesis that slow waves post-stroke are linked to a breakdown of causal interactions and signal complexity associated with a loss of motor function.

## MATERIAL AND METHODS

### Subjects

Forty patients with unilateral upper limb deficit ranging from mild to very severe motor deficits due to a first-ever stroke were recruited from the Department of Neurology, University Hospital of Cologne. Table 1 and the Supplementary Material I provide further details on the patients’ characteristics and inclusion and exclusion criteria [21].

**Table 1.** Patient characteristics.

| Patient No. | Group | Slow wave | Handed-ness | Lesion-side | Lesion location | Days post-stroke |  | MEP[μV] |  | NIHSS |  |  | Rel. grip strength[%] |  | ARAT |  |
| --- | --- | --- | --- | --- | --- | --- | --- | --- | --- | --- | --- | --- | --- | --- | --- | --- |
|  |  |  |  |  |  | Session I | Session II | Session I | Session II | Admission | Session I | Session II | Session I | Session II | Session I | Session II |
| 1 | I | x | R | R | Striato-capsular | 5 | 160 | 0.00 |  | 7 | 9 | 7 | 0.0 | 0.0 | 0 | 0 |
| 2 | I | o | R | R | partial MCA | 5 | 109 | 0.00 | 0.00 | 10 | 7 | 1 | 0.0 | 14.1 | 4 | 50 |
| 3 | I | o | R | R | Lenticulo-striate, parieto-occipital | 6 | 125 | 0.00 | 0.00 | 9 | 10 | 9 | 0.0 | 0.0 | 0 | 0 |
| 4 | I | o | R | R | Lenticulo-striate | 7 | 111 | 0.00 | 0.06 | 11 | 8 | 2 | 19.3 | 47.6 | 3 | 57 |
| 5 | I | o | R | L | pons | 5 |  | 0.00 |  | 15 | 12 |  | 0.0 |  | 0 |  |
| 7 | I | x | L | L | basal ganglia | 11 | 119 | 0.00 | 0.00 | 7 | 7 | 3 | 2.3 | 13.9 | 3 | 7 |
| 9 | I | x | R | L | partial MCA | 9 | 123 | 0.00 | 0.12 | 3 | 12 | 6 | 0.0 | 0.0 | 0 | 0 |
| 17 | I | o | R | L | pons | 6 | 175 | 0.00 | 0.00 | 8 | 12 | 7 | 0.0 | 0.0 | 0 | 0 |
| 18 | I | o | R | L | corona radiata | 7 | 126 | 0.00 | 0.11 | 3 | 10 | 2 | 0.0 | 63.5 | 0 | 20 |
| 19 | I | x | R | R | corona radiata | 2 | 125 | 0.00 | 0.00 | 3 | 10 | 6 | 0.0 | 0.0 | 0 | 3 |
| 22 | I | x | L | R | basal ganglia | 7 | 153 | 0.00 |  | 14 | 11 | 10 | 0.0 | 0.0 | 0 | 0 |
| 23 | I | o | R | L | partial MCA | 12 | 167 | 0.00 | 0.00 | 18 | 14 | 9 | 0.0 | 17.3 | 0 | 4 |
| 26 | I | o | R | L | internal capsule | 7 | 116 | 0.00 | 0.04 | 11 | 7 | 5 | 29.3 | 26.9 | 3 | 38 |
| 27 | I | o | R | R | medulla oblongata | 7 |  | 0.11 |  | 7 | 7 |  | 0.0 |  | 3 |  |
| 28 | I | x | R | L | Lenticulo-striate | 11 | 116 | 0.00 | 0.00 | 36 | 11 | 9 | 0.0 | 0.0 | 0 | 0 |
| 30 | I | o | R | R | partial MCA | 8 | 161 | 0.00 | 0.00 | 15 | 10 | 2 | 0.0 | 11.1 | 0 | 24 |
| 32 | I | x | R | L | MCA | 2 | 139 | 0.00 | 0.00 | 12 | 10 | 10 | 0.0 | 0.0 | 0 | 0 |
| 33 | I | x | R | R | MCA | 7 | 115 | 0.09 | 0.11 | 12 | 11 | 6 | 0.0 | 16.9 | 0 | 31 |
| 35 | I | x | R | R | partial MCA | 7 | 111 | 0.07 | 0.39 | 16 | 9 | 4 | 0.0 | 0.0 | 0 | 3 |
| 36 | I | o | R | R | partial MCA, internal capsule | 5 | 183 | 0.14 | 1.23 | 11 | 9 | 3 | 0.0 | 75.4 | 3 | 57 |
| 37 | I | o | R | L | corona radiata | 8 |  | 0.00 |  | 13 | 12 |  | 0.0 |  | 0 |  |
| 38 | I | x | R | L | corona radiata | 6 | 169 | 0.00 | 0.00 | 12 | 11 | 5 | 0.0 | 0.0 | 0 | 13 |
| 39 | I | o | R | L | Lenticulo-striate | 4 | 173 | 0.00 | 0.09 | 5 | 8 | 4 | 0.0 | 20.5 | 3 | 35 |
| 40 | I | o | R | R | corona radiata | 14 | 182 | 0.00 |  | 16 | 10 | 6 | 0.0 | 0.0 | 0 | 0 |
| 6 | II | o | R | R | partial MCA | 6 | 123 | 0.00 | 0.12 | 4 | 5 | 3 | 48.3 | 59.9 | 27 | 51 |
| 8 | II | o | L | L | internal capsule | 5 | 127 | 0.19 | 0.47 | 2 | 1 | 1 | 58.3 | 38.8 | 54 | 53 |
| 10 | II | o | R | R | Lenticulo-striate | 5 | 121 | 3.61 | 0.56 | 1 | 1 | 0 | 88.8 | 70.3 | 54 | 57 |
| 11 | II | o | R | R | MCA | 7 | 130 | 0.55 | 0.14 | 11 | 2 | 0 | 85.9 | 100.0 | 55 | 57 |
| 12 | II | o | R | R | pons | 9 | 110 | 0.61 | 1.83 | 6 | 4 | 0 | 94.7 | 91.3 | 48 | 57 |
| 13 | II | o | R | R | partial MCA | 2 | 170 | 2.47 | 1.53 | 7 | 4 | 0 | 79.5 | 86.1 | 45 | 51 |
| 14 | II | o | L | L | centrum semiovale | 6 |  | 0.16 |  | 1 | 1 |  | 70.4 |  | 53 |  |
| 15 | II | o | R | R | partial MCA | 9 | 149 | 0.28 | 0.17 | 5 | 4 | 0 | 48.7 | 92.1 | 48 | 57 |
| 16 | II | o | L | L | basal ganglia | 5 | 115 | 0.00 | 0.16 | 7 | 5 | 2 | 23.8 | 77.9 | 47 | 55 |
| 20 | II | o | R | R | partial MCA | 10 | 99 | 0.91 | 0.78 | 4 | 2 | 2 | 80.0 | 96.0 | 53 | 54 |
| 21 | II | o | R | L | internal capsule | 7 | 183 | 0.24 | 0.11 | 1 | 5 | 1 | 83.3 | 58.3 | 31 | 54 |
| 24 | II | o | R | R | partial MCA | 6 | 119 | 0.37 | 1.04 | 8 | 4 | 2 | 67.2 | 74.8 | 52 | 57 |
| 25 | II | o | R | L | partial MCA | 4 | 151 | 0.44 | 0.96 | 14 | 8 | 2 | 65.8 | 93.8 | 22 | 56 |
| 29 | II | o | R | L | partial MCA | 7 | 110 | 0.08 | 0.07 | 6 | 5 | 2 | 89.5 | 101.4 | 47 | 51 |
| 31 | II | o | R | R | thalamus | 7 | 177 | 1.66 | 2.87 | 19 | 7 | 3 | 78.8 | 99.3 | 44 | 55 |
| 34 | II | o | R | R | partial MCA | 2 |  | 0.12 |  | 8 | 3 |  | 43.5 |  | 39 |  |
| MEAN |  |  |  |  |  | 6.6 | 138.3 | 0.30 | 0.40 | 9.5 | 7.5 | 3.8 | 28.9 | 42.6 | 18.5 | 32.6 |
| SD |  |  |  |  |  | 2.7 | 26.6 | 0.72 | 0.66 | 6.5 | 3.6 | 3.1 | 35.9 | 38.9 | 22.8 | 24.4 |

(Group I: severely affected patients, Group II: mildly and moderately affected patients; please note that patients who were assigned to the group of severely affected patients are marked in blue and patients additionally featuring sleep-like responses are marked in red)

Thirty-five patients could be re-assessed behaviorally, and twenty-eight patients also participated in the TMS-EEG follow-up visit at least 3 months later (138.3±26.6; 99-183 days). Fifteen age-matched healthy participants served as a control group. Of note, parts of the original data, i.e., data from 28 patients obtained in the first two weeks post-stroke, and the healthy controls’ data, were previously included in [21]. Importantly, there is no relevant overlap between the analyses presented here and those published before, as the former was neither on local sleep nor on longitudinal TMS-EEG changes. Hence, all analyses presented here are entirely novel.

All participants provided informed written consent before entering the study. All aspects were approved by the ethics committee of the Medical faculty of the University Cologne (file no. 17-244) and carried out in accordance with the Declaration of Helsinki (October 2013).

### Experimental procedure and data collection

We assessed three motor and clinical parameters within (i) the first two weeks and (ii) more than three months post-stroke: i) NIHSS, ii) ARAT [24], and iii) relative grip strength [21].

TMS was performed using a Magstim Super Rapid^2^ stimulator (The Magstim Co. Ltd, Whitland, United Kingdom) equipped with a Magstim 70mm figure-of-eight Alpha Film Coil. TMS was applied over the ipsilesional primary motor cortex (M1). The ‘motor hotspot’ for M1 was defined as the coil position eliciting MEPs of the highest amplitude of the first interosseous (FDI) muscle in response to a TMS-pulse applied tangentially to the skull in a 45° posterior-anterior direction. MEP recordings were obtained by Ag/AgCl surface electrodes (Tyco Healthcare) in a belly-to-tendon montage. The EMG signal was amplified, filtered (0.5Hz high pass and 30-300Hz bandpass), and digitized using a Powerlab 26T device and the LabChart software package version 8.0 (AD Instruments, Sydney, Australia). The RMT was defined using an algorithm provided by the TMS Motor Threshold Assessment Tool (MTAT) 2.0 (http://www.clinicalresearcher.org/software.html) [25]. In case no MEPs could not be evoked upon ipsilesional M1 stimulation even with maximal stimulator output, we used the contralesional RMT as a reference for the individual threshold (n=23) in order to avoid excessively high stimulation intensities at the structurally intact motor cortex due to lesion-induced subcortical disruption of the corticospinal tract [21]. Using this procedure warranted that the factual average stimulation intensity did not significantly differ between patients (65.2±11.6% MSO) and healthy individuals (62.0±14.8% MSO) neither in the first weeks post-stroke (p=0.4) nor after more than three months (57.9±9.3% MSO, p=0.3). Moreover, in case of MEP-negativity, we used anatomical landmarks, i.e., the hand knob, to define the ipsilesional motor hotspot. Otherwise, the cortical excitability of ipsilesional M1 was assessed by MEP amplitudes evoking ten MEPs with 110% of individual RMT randomly jittered at 6.0-8.5s. The position of the coil was tracked and recorded using a neuronavigation system throughout the experiment (BrainSight V.2.0.7; Rogue Research Ltd; Montreal, Canada). For this purpose, the head of the participants was co-registered with an individual anatomical MR image.

During the EEG recordings, 100-150 trials of single TMS-pulses in each session were applied to ipsilesional M1 with an inter-trial interval randomly jittered between 6.5-8.0s with 80% RMT [21,26–28].

TMS-EEG was recorded using a TMS-compatible 64-channel EEG-system (BrainAmp DC, BrainProducts GmbH, Gilching, Germany). Scalp EEG was recorded by 62 TMS-compatible Ag/AgCl sintered C-ring electrodes mounted on an elastic electrode cap (EasyCap-Fast’n Easy 64Ch) following the standard layout and the international 10-20 system arrangement. The EEG-signals were sampled at a frequency of 5kHz with a resolution of 0.1µV, and filtered with high-pass 0.1Hz and low-pass 1kHz. The impedance of all electrodes was constantly kept below 5kΩ. Preparations and utilized hardware ensured high-quality recordings and minimization of artefacts (Supplementary Material II for details on TMS-EEG recordings).

### Data analysis

Data analysis was performed using Matlab (The MathWorks, Massachusetts, USA). First, TMS-EEG recordings were visually inspected to reject single trials and channels containing noise or muscle artifacts [9,19,21]. Recordings with either less than 90 artifact-free trials or more than 10 poor channels were excluded from further analysis (number of artifact-free single trials: patients: 99.4±10.2 SD; healthy controls: 104.1±19.8 SD). The TMS artifact between −2ms and 8ms relative to TMS pulse onset was removed. Missing data were replaced with baseline [21,29]. Subsequently, data were detrended, band-pass and band-stop filtered (1-60Hz; 49-51Hz; Butterworth 3^rd^ order), down-sampled at 625Hz, and segmented in time windows of ±1000ms around the TMS pulse artifacts [19,21] Poor channels were spherically interpolated using the respective EEGLAB function (https://sccn.ucsd.edu/eeglab/) [30].

Trials were average re-referenced and baseline corrected [19]. Finally, independent component analysis (ICA) was applied to remove residual TMS-related artifacts and ocular or muscle artifacts [31]. Of note, data processing was carried out identically to [9,18,19,21].

Based on previous findings and to comparatively characterize the evolution of TMS-EEG alterations longitudinally over time post-stroke [21], we calculated the Local Mean Field Power (LMFP) of ipsilesional M1, quantifying the evoked electric field as a function of time for the channels closest to the stimulation site stimulation (FC3-FC1-C3-C1/FC4-FC2-C4-C2 for ipsilesional M1). LMFP time series significantly different from baseline [26,32] were used to compute the integral between +8 and +350ms post-stimulus and entered into group analysis.

Additionally, spectral features were evaluated by computing the event-related spectral perturbation (ERSP) between 5 and 50Hz after time-frequency decomposition using the Morlet wavelet transform [26,28]. For subsequent analyses, similar to LMFP, only significant ERSP values surviving the bootstrap-based statistics were considered to extract the frequency with the maximum power, i.e., the natural frequency [28], of the TMS-evoked response for ipsilesional M1.

To explicitly address the phenomenon of sleep-like slow waves, we further assessed the amplitude of TMS-evoked low-frequency components [7,9]. Therefore, single trials were low-pass-filtered (<4Hz), averaged, and rectified, before for each channel the maximum slow wave amplitude was computed for the time window of 8-350ms post-stimulus.

Besides, with explicit regard to cortical off-periods associated with slow waves, a manifestation of cortical bistability, the suppression of high-frequency EEG power (>20Hz) was assessed [7,9]. Accordingly, the integral of high-frequency power between the time period for which desynchronization is expected (100-350ms after TMS onset) for the ipsilesional M1 was determined [9,18].

Since these phenomena are thought to hinder the formation of complex network interactions, we quantified TMS-induced local causal interactions by means of the significant time course of the phase-locking factor (PLF) averaged over the four channels closest to the stimulation site (FC3-FC1-C3-C1/FC4-FC2-C4-C2 for ipsilesional M1) [7,9]. The PLF, calculated for every single electrode as the absolute value of the average of the Hilbert Transform across trials, is suggested to be a proxy of connectivity evaluating the instantaneous phase difference assuming that functionally connected areas should be synchronized across trials. The latest significant PLF time point was identified and averaged over the four channels. Finally, to assess the complexity of global causal interactions, we calculated the perturbational complexity index (PCI), capturing the deterministic spatiotemporal patterns of neural activation in response to a direct perturbation reflecting the joint ability of integration and differentiation [19,33]. Comprehensive details on the calculation of the former TMS-EEG parameters could be found in the Supplementary Material III.

### Lesion Quantification

For all stroke patients, lesion maps were created and processed using standard procedures (Supplementary Material IV). We subsequently calculated the lesion volume, the probabilistic volume of the overlap with the cortical spinal tract (CST), and a weighted overlap accounting for the narrowing of the descending CST [34,35]. However, as stroke is conceptualized as a network disorder, we quantified the effects of the focal lesion on the brain’s connectome using the Lesion Quantification Toolkit (https://wustl.box.com/v/LesionQuantificationToolkit [36]; Supplementary Material IV). Please note that lesion data were particularly used to derive additional information about TMS-EEG characteristics and to differentiate clinically (severely vs. mildly to moderately affected) or neurophysiologically, i.e., by means of TMS-EEG responses (slow wave vs. complex response), defined subgroups. To create subgroups based in lesion location was beyond the scope of the present work.

### Statistical analysis

Statistical analyses were performed using the software SPSS (v28, IBM). According to Shapiro-Wilk tests to decide on parametric or non-parametric statistics, significant differences were evaluated using independent t-tests or Mann-Whitney U tests to test between-group comparisons and paired t-tests or Wilcoxon tests for within-group comparisons between the two sessions (p<0.05). Tests were always two-sided, post-hoc tests were Bonferroni-corrected for multiple comparisons. Motor and recovery scores were used as a principal component analysis (PCA)-generated composite score [21,37]. To derive the motor composite score for the first days after stroke, we entered the individual (i) ARAT score and (ii) relative grip strength of the first session into a PCA, in which the first component represented the aggregated deficit of the upper limb. The same approach was pursued for the composite score of the residual deficit utilizing the individual scores of the second assessment after more than three months. To generate the composite recovery score, we calculated the relative differences between the first session and the follow-up for each motor parameter and subsequently entered the z-standardized difference scores into a PCA. Notably, the NIHSS score was not included, as it rather reflects the global neurological impairment.

To test for the clinical relevance of TMS-EEG alterations, also in comparison the known predictors for motor outcome, i.e., MEP-status or initial impairment, linear regression analyses with the composite recovery score as the dependent variable and initial motor composite score and MEP amplitudes or TMS-EEG parameters as independent variables were computed.

## RESULTS

### Motor reorganization is paralleled by a reinstatement of signal complexity

After 3-6 months post-stroke, the majority of patients experienced substantial recovery (NIHSS: p<0.001, t_(34)_=10.13; motor composite score: p<0.001, t_(34)_=-4.80) (Figure 1).

**Figure 1.**
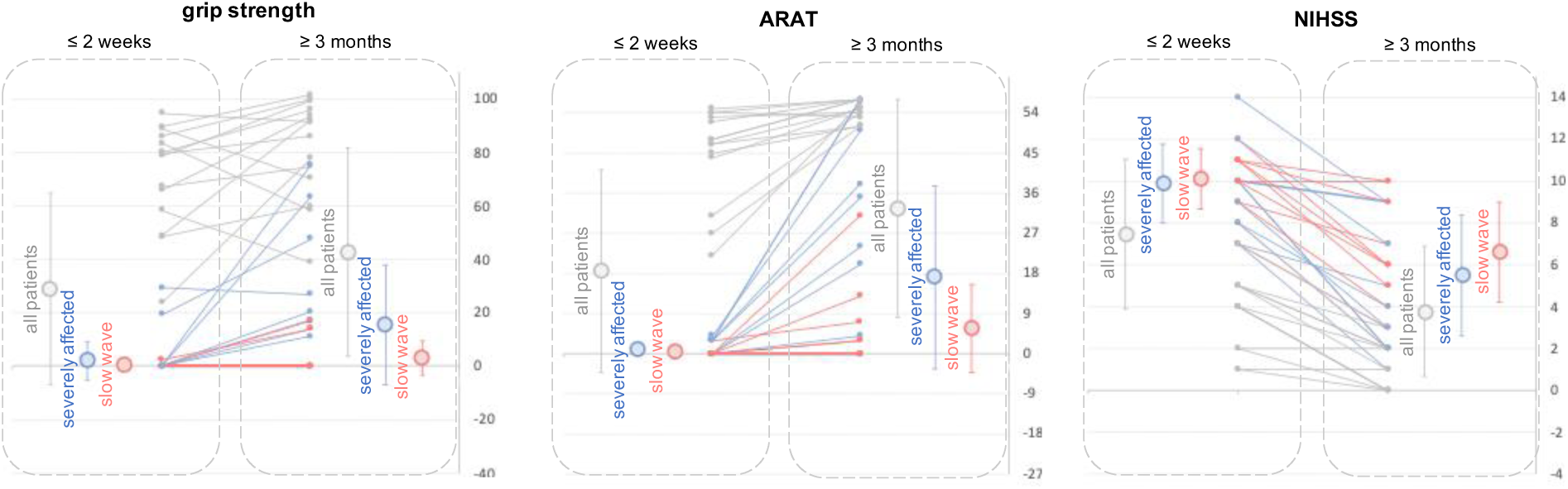
Longitudinal clinical data. Individual and mean average recovery data of the motor scores (grip strength, ARAT) and the NIHSS (error bars indicate standard deviation). Subgroups of patients are color-coded (Table 1). Please note that while all patients and also the subgroup of patients with initially more severe motor deficits (n=24; k-means clustering by the motor scores) showed substantial recovery as indexed by a decrease in the NIHSS and an increase in motor performance after more than three months post-stroke, the subgroup with sleep-like TMS-EEG responses early post-stroke only depicted an improvement of the general neurological deficit but not of specific motor scores.

Concurrently, we observed a reinstatement of complex TMS-evoked EEG-responses with a decrease in the initially elevated amplitudes and an increase in long-lasting oscillations. Source reconstruction confirmed that, in parallel to recovery of function, TMS-induced activity evolved to distributed patterns particularly involving both hemispheres, approximating those of healthy controls (Figures 2A-C).

**Figure 2.**
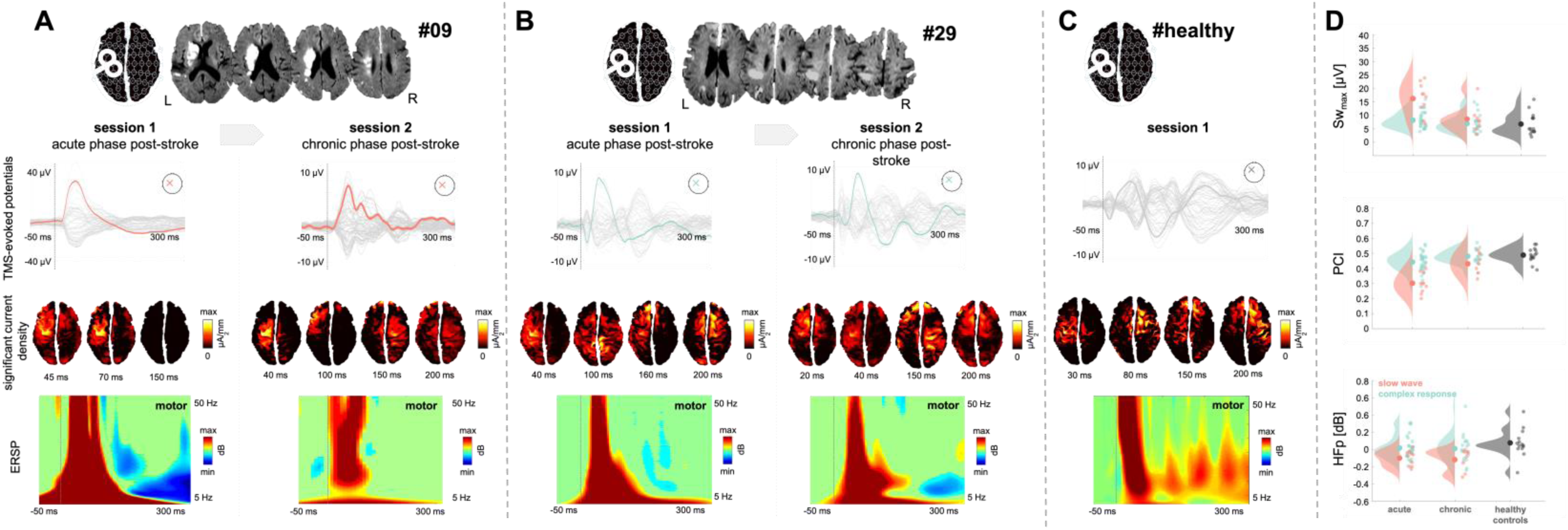
Local sleep-like cortical off-periods over ipsilesional M1 impair global complexity and vanish with the motor reorganization. Representative results of individual data (A) with a clear slow-wave response early post-stroke in contrast to (B) a complex response and (C) a healthy individual. Top row: Individual lesion location and clinical information. Second row: Evolution of average TMS-evoked potentials over time post-stroke (all channels superimposed in grey, stimulation electrode C3 colored trace). Third row: Significant current density cortical maps are shown at selected time points for each patient. Fourth row: Event-related spectral perturbation (ERSP) for the ipsilesional motor cortex. Please note for patient A in the first session that the slow wave response is associated with high-frequency power decrease in line four and a stationarity of the signal in line 3. Moreover, with time post-stroke the disappearance of slow waves and increase of signal complexity in (A), particularly depicting a reinstatement of global connections after 3 to 6 six months post-stroke. Raincloud plots (D) and individual values of the slow wave amplitude (SWmax), perturbational complexity index (PCI), and suppression of high-frequency power (HFp) for the 1^st^ (acute) and 2^nd^ (chronic) session of the slow-wave response group (red) and the rest of the patients (mint) and healthy controls (grey).

Correspondingly, the LMFP of ipsilesional M1, which was increased early post-stroke (p=0.006, t_(54)_=3.3), significantly decreased during reorganization (p=0.015, t_(57.5)_=2.9) and approached levels of healthy participants (p=0.47) (Figure 3).

**Figure 3.**
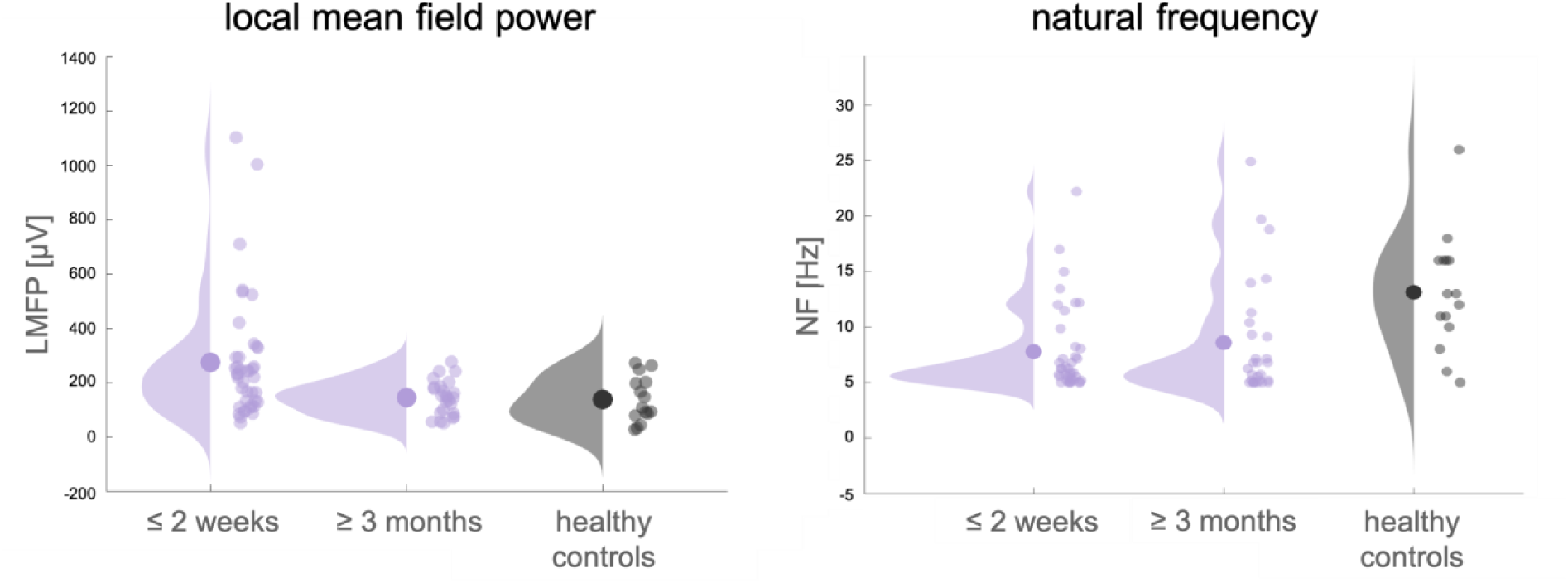
TMS-EEG response changes with motor reorganization. Raincloud plots with individual data of the development of the LMFP and natural frequency throughout motor reorganization (lilac) compared to healthy participants (grey). While early after stroke, local mean field power (LMFP) of the ipsilesional motor cortex was particularly increased compared control subjects, LMFP subsequently decreased during reorganization. TMS-evoked oscillations, i.e., natural frequency (NF), was altered with a stroke-associated slowing both within the first weeks after stroke and in the follow-up.

**Figure 4.**
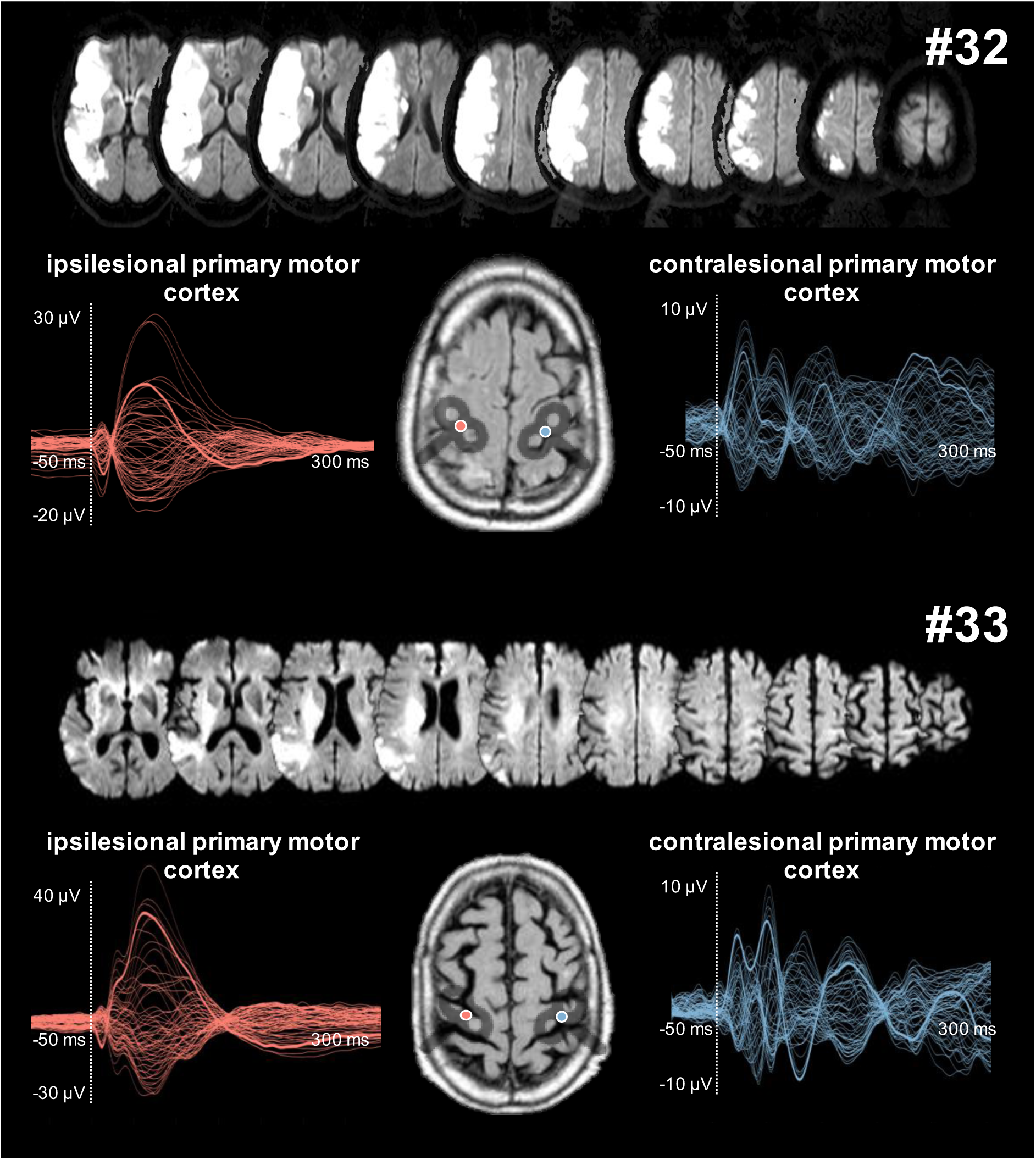
Ipsi- and contralesional stimulation. TMS-evoked sleep-like responses are a characteristic of the stimulation of the ipsilesional motor cortex but not of the stimulation of the contralesional motor cortex. In the unaffected hemisphere, the TMS-EEG responses markedly differed from the ipsilesional motor cortex, as they were composed of low-amplitude, fast-frequency oscillations. Hence, the phenomenon of sleep-like waves was specific to the lesioned hemisphere. Besides, we observed these responses in a broad spectrum of ischemic lesions ranging from smaller subcortical to larger defects involving subcortical and cortical structures (Table 1).

By contrast, alterations in the time-frequency domain with a slowing of TMS-evoked oscillations in ipsilesional M1 early post-stroke (p=0.002, Z=-3.27) remained altered also 3-6 months post-stroke compared to healthy controls (p=0.01, Z=-2.79) with basically no difference compared to the early stroke phase (p=0.85), indicating a sustained disturbance of cortical oscillatory properties over time post-stroke (Figure 3).

We next aimed at quantifying changes in network interactions: Accordingly, early after stroke, the duration of TMS-evoked effects on local interactions (PLF_t_) was significantly shorter in patients (p=0.016, t_(54)_=-2.78) and showed no significant difference after three months post-stroke (p=0.67), implying that local TMS-induced neural interactions were still impaired in this later phase.

Furthermore, we subsequently quantified the alterations of global signal processing and transmission, as assessed by PCI [33]. Mean PCI-values of patients early post-stroke were significantly lower compared to healthy controls (p<0.001, t_(44)_=-4.32). However, with motor reorganization, we observed a restoration of global signal complexity over ipsilesional M1, as PCI increased with time post-stroke (p=0.006, t_(63.3)_=-3.21) toward values of healthy participants (p=0.6).

Overall, motor reorganization was accompanied by a restoration of long-range transmission as indexed by an increase in global complexity alongside recovery, while the local processing of the TMS-induced activity was still altered in the chronic phase post-stroke.

### Complexity as a marker for motor recovery

Crucially, the persistent TMS-EEG signal alterations more than three months post-stroke had to be considered functionally significant: The residual motor deficit was linked to the duration of causal effects on local cortical interactions (PLF_t_) assessed in the second session (r=0.59, p<0.001), indicating that more pronounced persistent local signal alterations were accompanied by stronger residual deficits (Supplementary Figure 1B). However, estimating the potential for recovery and, thereby, the permanent deficit very early post-stroke is particularly critical. Thus, we used a stepwise multiple linear regression to elucidate information about motor recovery based on (i) the initial motor deficit, (ii) MEP-amplitudes [21], and (iii) the TMS-EEG parameters assessed early post-stroke. Here, the initial motor deficit and the PCI were most indicative of motor recovery (R^2^=0.49, adjusted R^2^=0.44, F_(2,22)_=9.50, p=0.002) with PCI explaining 21% of unique variance (Supplementary Figure 1A). Importantly, in the clinically and electrophysiologically homogenous, i.e., MEP negative, subgroup of initially severely affected patients (n=20), in whom established predictors of post-stroke recovery such as MEP-amplitude or initial impairment fail to differentiate between levels of recovery [21], the same model, precisely also containing the acute motor impairment, identified the initial PCI as the sole predictor, explaining 65% of the variance of motor recovery (adjusted R^2^=0.60, F_(1,9)_=12.01, p=0.016) (Supplementary Figure 1C).

### Cortical off-periods that impair global complexity vanish with time post-stroke

In a subgroup of the patients (25%, Table 1), the stimulation of ipsilesional M1 early post-stroke evoked a simple biphasic, i.e., positive-negative, slow-wave response associated with an initial high-amplitude, broad-band activation followed by a suppression of high-frequency power (Figure 2A). These responses occurred exclusively in patients with initially severe impairments, and explicitly differed from the response profiles of healthy individuals in whom slow waves were always absent. By contrast, the subgroup of patients featuring a typical slow-wave response particularly exhibited higher low-frequency amplitudes (<4Hz; SW_max_) (healthy individuals:p=0.002, Z=-3.0; between patients: p=0.002, Z=-2.94) (Figure 2D). Moreover, these slow waves were associated with a significant suppression of high-frequency EEG power (>20Hz; HFp), that was exclusively detectable in the slow-wave response group (healthy individuals: p=0.005, Z=-2.72; between patients: p=0.03, Z=-2.16), complying with the criteria for sleep-like off-periods [7,9].

This slow wave response early post-stroke remained stationary at the stimulation site in ipsilesional M1, as revealed by source analysis (Figure 2A) and also affected the interactions with other brain regions. In this context, the duration of TMS-effects on local connections was significantly shorter in the patient group with slow wave responses (healthy individuals: p<0.001, t_(18)_=-4.02; between patients: p=0.014, t_(38.6)_=-2.58). With regard to alterations of global interactions, the mean PCI-value of the slow-wave response subgroup was around the empirical threshold reported for consciousness (0.31±0.07; PCI-threshold: 0.31 [19]). Notably, the PCI-values of seven patients were invariably lower than this threshold (Figure 2D) and hence comparable to ranges found for unconscious conditions [19]. In line with that, a reduction of global signal complexity was particularly evident for the patient group exhibiting slow waves (healthy individuals: p<0.001, t_(23)_=-7.50; between patients: p<0.001, t_(39)_=-5.51) (Figure 2C), indicating substantial disturbances of information processing and network communication.

In the follow-up assessment, contrary to the entire stroke sample, this subgroup did not show a significant improvement in motor performance (p=0.23), while improvements in other functional domains as captured by the NIHSS (p<0.001, t_(9)_=5.22) were also present (Figure 1).

Crucially, slow waves observed in the first weeks vanished in all patients in the follow-up measurement. As we kept the TMS-target consistent across sessions, these changes do not only reflect processes engaged in reorganization but also disclose information about the putative underlying substrates of slow waves in stroke.

Likewise, we found a significant decrease in low-frequency amplitudes from the first weeks post-stroke to the three months follow-up (p=0.029, Z=-2.20) and, consistent with this, no significant difference anymore (healthy controls: p=0.39, between patients: p=0.35) (Figure 2A-D). By contrast, suppression of high-frequency EEG-power remained evident in the follow-up assessment (healthy individuals: p=0.01, Z=-2.71; between patients: p=0.033, Z=-2.14; time post-stroke: p=0.97) (Figure 2D).

While source reconstruction suggested a restoration of signal transfer within the functional network of ipsilesional M1 three months post-stroke, the duration of TMS-induced deterministic effects near the stimulation site (PLF_t_) did not change throughout motor reorganization for the slow wave response group (p=0.69). However, with motor system reorganization, we observed an increase in global signal complexity for ipsilesional M1-stimulation from the post-acute to the later phase (p=0.004, t_(15)_=-3.42). Of note, at the follow-up, all PCI-values reached levels over and above the empirical threshold reported for consciousness [19] (Figure 2D).

### Correlates of cortical off-periods in stroke

All patients with a characteristic slow wave response were assigned to the group of severely affected patients and presented a more pronounced functional deficit in the first weeks post-stroke compared to all other patients (NIHSS: p<0.001, t_(36.8)_=4.4; motor score: p<0.001, t_(30.2)_=-5.75). However, within the severely affected subgroup, there was no difference in impairment for patients with and without slow waves (NIHSS: p=0.53; motor score: p=0.36) (Figure 1). Thus, slow waves upon ipsilesional M1-stimulation arise with severe impairment but are not merely an expression of a stronger deficit as they rather differentiate between clinically homogeneous patients. In fact, analyzing the amount of recovery and the residual motor deficit revealed that the slow-wave response group sharply demonstrated an attenuated functional recovery and, thereby, an increased permanent deficit in the follow-up visit compared to both the entire patient cohort but also the severely affected group (recovery: all: p=0.013, t_(32.3)_=-2.62; severely affected: p=0.015, t_(13.1)_=-2.8; residual deficit: all: p<0.001, t_(32.3)_=-7.61; severely affected: p=0.012, t_(12.9)_=-2.94) (Figure 1). Crucially, the fact that the slow wave subgroup and the other patients within the group of severely affected did not differ with respect to the initial degree of impairment particularly emphasizes the predictive potential of slow waves and cortical off-periods for recovery and residual function in otherwise undistinguishable clinical phenotypes.

### The hypothesis of disconnection

The highest overlap of the individual lesions was located at the level of the basal ganglia, i.e., in the posterior limb of the internal capsule, pallidum, and putamen (Figure 5A).

**Figure 5.**
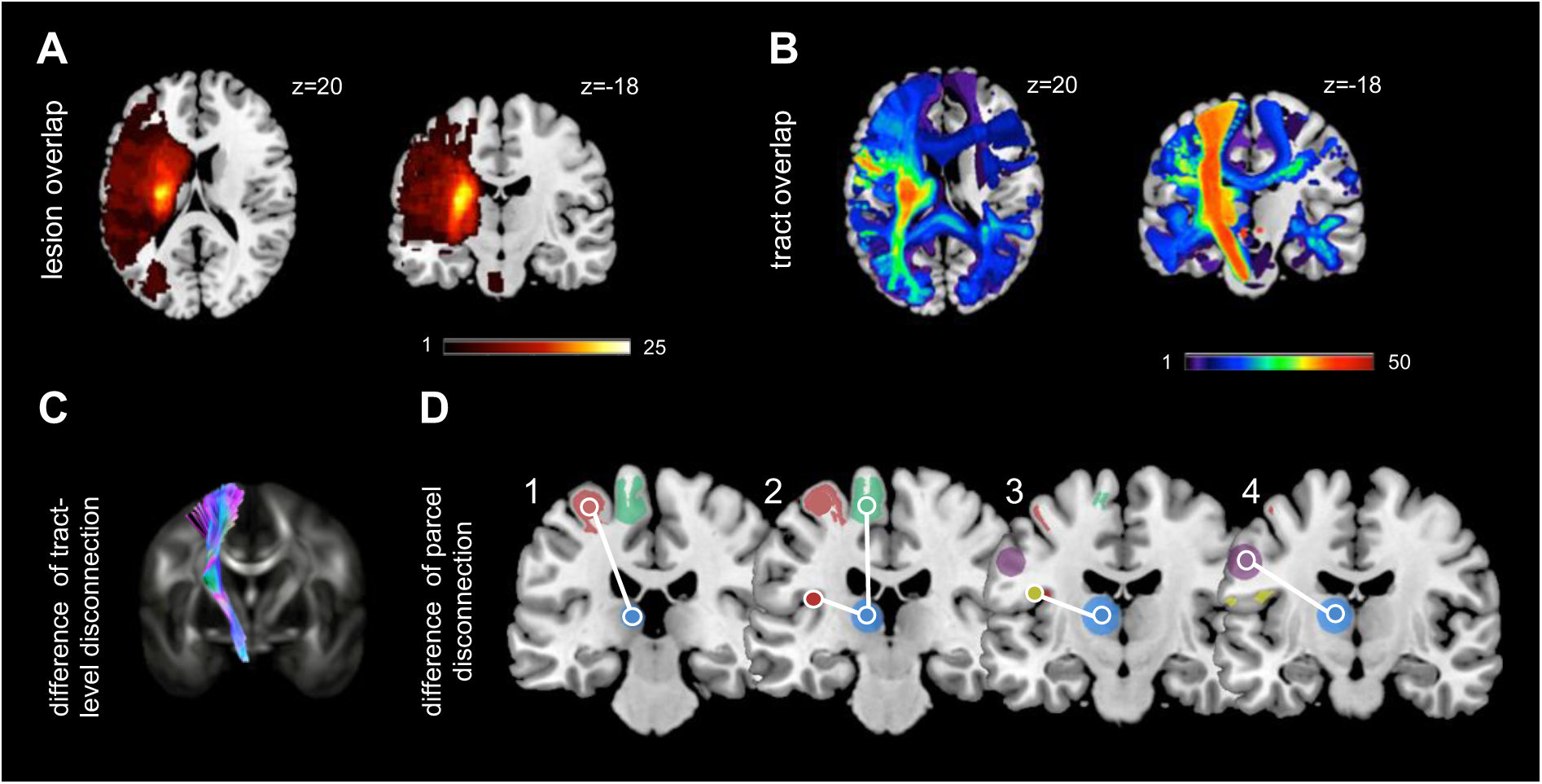
Lesion and disconnection maps. Lesion overlap (A) of stroke patients, based on T2 images. Overlap of lesion-induced white matter tract disconnections (B) and significant differences of the subgroup with sleep-like responses compared to all patients and severely affected in fiber tract disconnection (pedunculopontine tegmental nucleus (PPT) (C) and parcel disconnection of somatomotor parcels (1: SomMot A1 (red); 2: SomMot A2 (green) and SomMot BS2 1 (red); 3: SomMot BS2 2 (yellow); 4: SomMot BCent (lilac)) and caudate nucleus (blue).

We neither found a difference in lesion volume nor lesion-induced damage to a particular brain area in patients presenting with or without slow-wave responses (Figure 5; Supplementary Material V), indicating that slow waves and cortical off-periods post-stroke are not merely a consequence of lesion load.

However, when analyzing the amount of fiber tract disconnection, i.e., the severity of lesion-induced tract-level white matter disconnection [36], patients presenting with slow wave responses featured lesions in the ipsilesional pedunculopontine tegmental nucleus (PPT), which is a component of the ascending activating systems located in the pontomesencephalic reticular formation (all: p<0.001, t_(37)_=4.77; severely affected: p=0.008, t_(19.5)_=2.96) (Figure 5C). Moreover, when quantifying the disconnection severity in the structural connectome [36], we found more severe disconnections in the ipsilesional hemisphere in patients with slow waves between somatomotor areas and ipsilesional caudate nucleus (Figure 5D).

## DISCUSSION

We provide an extensive and longitudinal analysis of TMS-induced signal alterations and electrophysiological mechanisms occurring in the motor system early post-stroke and alongside neural reorganization. We show that not only in the early post-acute phase but also after more than three months the TMS-EEG response depicts both information about reorganization and signal alterations associated with persistent functional deficits, underlining the significance throughout the entire post-stroke period. Complementing and expanding previous findings [18,21], we demonstrate that in a fraction of patients with severe motor deficits, TMS applied to ipsilesional M1 evoked a slow-wave response associated with a cortical off-period and underpinned by a deterioration of global signal complexity and disruption of the TMS-induced formation of causal network interactions. However, tracking these signal alterations and the course of motor recovery during reorganization revealed that slow waves were only observed in the first days and disappeared in the later phase. Crucially, their occurrence early post-stroke was an indicator of a poor motor outcome.

Hence, we not only establish a link between the pathophysiology of acute stroke and cortical bistability, a neuronal mechanism that has been precisely characterized in animal models and NREM sleep and subsequently associated with states of unconsciousness, but put it into a behaviorally and clinically meaningful perspective. Thus, our data reveal novel information about the neural underpinnings of slow waves, thereby providing crucial insights into the neurobiological mechanisms driving reorganization and recovery of function.

The classical notion of lateralized slowing in the cortical vicinity of lesions has been derived almost a century ago in early EEG recordings [38,39]. However, the pathophysiological understanding of stroke-related EEG slowing is still limited. By contrast, the mechanisms of slow waves in physiological conditions have been comprehensively identified in in-vitro, in-vivo, and in-silico models. Accordingly, large slow waves associated with transient suppressions of high-frequency activity, i.e., off-periods, reflect cortical bistability and are a hallmark NREM sleep [7,40]. The phenomenon of cortical bistability entails a profound membrane hyperpolarization after a transient activity increase [10]. More precisely, after an initial activation, e.g., triggered by the external TMS-perturbation, neurons rapidly plunge into a silent down-state that disrupts the emergence of complex long-range responses and sustained interactions usually observed during wakefulness [9,10,41]. It has been suggested that this phenomenon is influenced by enhanced adaptation mechanisms such as activity-dependent potassium currents, decreased modulation from activating systems, and increased GABAergic inhibition [11–13,42]. While cortical bistability represents a mode of physiological activity during sleep [43], slow waves and cortical bistability could also be triggered by pathological alterations inducing shifts in the balance between inhibition and excitation, or white matter lesions [9,44–47]. In line with this notion, recent work using TMS-EEG has confirmed the neuronal principles of slow wave, cortical off-periods, and bistability, indistinguishable from sleep slow waves, in brain injury and disorders of consciousness [9]. Two TMS-EEG studies have lately established that slow waves can intrude into wakefulness following brain lesions by revealing the occurrence of local slow waves in conscious, awake stroke patients [18,21], and raising the question of whether the neural mechanisms identified for cortical bistability in NREM sleep and unresponsive wakefulness might also apply here.

Of note, while the former two conditions, i.e., sleep and unconsciousness, are characterized by sleep-like waves and cortical off-periods as ubiquitous cortical features irrespective of the stimulation site, the occurrence of slow waves in stroke has so far been shown only locally and restricted to the ipsilesional hemisphere [18,21]. Extending previous findings, we show that the local perturbation of ipsilesional M1 is associated with a global deterioration of signal complexity and long-range interactions, paralleling results in sleep and unconsciousness [9,19].

There is compelling evidence, in-vivo and in-vitro, on the existence of locally confined sleep phenomena at the neuronal level [48,49], which may clarify the phenomenon observed in stroke. Besides the classical concepts of sleep and wakefulness as discrete global states, cortical neurons can briefly fall into off-periods accompanied by the occurrence of slow waves as they do in sleep but locally restricted to a distinct area and fully awake with the characteristic low-voltage fast-oscillatory activity [6]. Thus, local sleep shares similar electrophysiological properties with those encountered in sleep but as a local process under local regulation coexisting with widespread features of wakefulness [48]. While we link slow waves over ipsilesional M1 to motor deficits, the analogy manifests even more strikingly, since the occurrence of local sleep in circumscribed regions has been associated with behavioral deficits functionally directly linked to the altered part of the cortex [5,6].

A further approach to reconcile the neural underpinnings of stroke and sleep lies in the notion of neuroplasticity. While consensus prevails that sleep, and in particular slow-wave sleep, is tightly connected to mechanisms of synaptic plasticity [22,50], ischemia triggers a cascade of cellular and biochemical processes resulting in the formation of new synapses and axonal sprouting governing neuroplasticity and restoration of function [1,51]. Of note, axonal sprouting post-stroke has been associated with low-frequency oscillations similar to slow-wave sleep [2,3]. Especially relevant in the context of plasticity, immune signaling molecules, particularly cytokines like interleukin-1 (IL-1) and tumor necrosis factor alpha (TNFα), are supposed to locally promote physiological sleep and are related to EEG slow waves [52–54]. Correspondingly, given that enhanced IL-1 and TNFα-levels have been observed post-stroke [55] and higher cytokines levels are linked to stroke severity [56,57], these data align with our findings of slow waves in severely affected patients early post-stroke, i.e., when cytokines are strongly upregulated.

The crucial question, however, lies in the functional relevance of slow waves and off-periods in stroke. While they might represent similar recovery processes as in physiological sleep, necessary for neuronal stabilization [58], and slow waves have been associated to axonal sprouting after stroke potentially supporting reorganization, our data rather imply that slow waves are rather associated with attenuated recovery processes and persistent disability. In other words, the neuronal mechanisms leading to slow-wave responses might not facilitate recovery of function but instead indicate a poor outcome. Although hypothetical, the functional significance of stroke-associated slow waves might also be subjected to temporal changes comparable to other processes such as GABAergic inhibition [59,60], potentially neuroprotective in the very acute phase but subsequently with subacute persistence facilitate maladaptive processes interfering with recovery. Along these lines, cortical bistability has been associated with a shift in the inhibition/excitation-balance towards excessive inhibition [11,44]. In parallel, increased inhibition has been found to arise locally post-stroke [15,61] and blocking increased tonic GABA-mediated inhibition in the peri-infarct tissue facilitated functional recovery at later stages post-stroke [15,59]. A speculative yet very interesting theory is that slow waves post-stroke may not only be an epiphenomenon of mechanisms leading to less successful recovery but might even constitute the direct manifestation of a vast inhibitory tone, potentially blocking functional reorganization and thereby at least in parts leading to a less favorable outcome.

Beyond, yet in context, the proposition of disconnection of brain regions due to a structural lesion warrants further consideration. The phenomena discussed above are likely to result from disconnection effects. A critical reduction of cortico-cortical connectivity has been suggested to alter the inhibition/excitation-balance [11,44]. Furthermore, isolated cortex preparations have been shown to generate slow activity, implying a cortical origin of slow waves and, thereby, underlining the notion that the lesion-induced disconnection and isolation of cortical regions might lead to slow waves [45,62,63]. Likewise, a cortical undercut - an animal model of cortical deafferentation - resulted in slow waves in the deafferented region, even in awake animals [47]. Therefore, white matter lesions not only affect output fibers of the cortex but also interrupt input fibers and thus yield a cortical deafferentation from ascending activating systems [20]. Accordingly, a recent study combining tractography analysis and TMS-EEG found local sleep-like responses selectively circumscribed to cortical sites whose afferents were interrupted by a subcortical focal lesion [64]. Besides the structural disconnection, lesions may also result in functional alterations enhancing the tendency of intact parts of the thalamocortical system to shift into silent down states, preventing a complex propagation [9,10,38].

In this context, a critical relevance of the thalamus has been postulated for the generation of fast-frequency oscillations [65] and slower rhythms, constantly found in stroke patients early and later post-stroke, likely reflect a hyperpolarized thalamocortical system, driven mainly by an increased inhibition and deficient neuromodulatory arousal systems. Moreover, the thalamus and thalamocortical connections in the affected hemisphere have been shown to play a crucial role in reorganization, neuroplasticity, and functional recovery post-stroke [66].

In addition, we linked slow waves and cortical off-periods to lesion-induced tract disconnection of fibers in the pedunculopontine tegmental (PPT) nucleus. The PPT, as part of the ascending reticular activating system, projects significant input to the thalamus, giving rise to affect cortical activity through thalamocortical connections, and is reciprocally connected to the basal ganglia [67]. Crucially, the PPT has been related to the generation of sleep-like waves and bistability in animal models [12,42], supporting our data. Along these lines, we have further linked slow wave responses in stroke to a disconnection of the structural connectome between somatomotor regions and the caudate nucleus. The latter is known to be involved in regulating cortical excitability by predominantly inhibitory modulation [68,69]. Since slow-wave sleep has been connected to a decrease in the activity of the caudate nucleus [70], interruption of its cortical projections may result in comparable excitability levels finding expression in sleep-like waves and cortical off-periods observed post-stroke.

### Limitations and Conclusion

However, the exact mechanistic underpinnings remain open, as invasive recordings in humans and direct evidences from animal models are yet missing. Moreover, our findings are currently limited to the stimulation of ipsilesional M1, which is the crucial region for motor reorganization, but certainly has to be understood within the network architecture of a broad motor system. Thus, systematically assessing other frontoparietal key motor areas known to play a role in motor control after stroke [71–73] constitutes an important future goal. Synoptically, our findings render the notion plausible that an ischemic lesion, affecting critical cortico-cortical and cortico-subcortical connections, might lead to decreased modulations of ascending activating systems and alterations in the balance between excitation and inhibition.

In this vein, the question arises whether neuromodulatory intervention may alleviate increased inhibition, push neurons beyond bistability, and, thus, promote recovery of function [74]. Although we only assessed two time points, rendering a dense monitoring to capture the temporal evolution of slow waves desirable, the phenomenon of slow waves as a putative signature of an increased inhibitory tone leading to dysfunctional reorganization may offer a new therapeutic strategy to improve the functional outcomes. In consequence, TMS-EEG might offer a personalized readout for targeting and titrating interventions aiming at restoring complex connectivity patterns.

## Data Availability

All data produced in the present study are available upon reasonable request to the authors.

## ACKNOWLEDGMENTS

GRF and CG are funded by the Deutsche Forschungsgemeinschaft (DFG, German Research Foundation) – Project-ID 431549029 – SFB 1451 (project C05). UZ is funded by the European Research Council (synergy grant ConnectToBrain, Grant agreement No. 810377) and the Federal Ministry of Education and Research (BMBF, project-ID 01KG2125). We are grateful to Sebastian Dern, Jana Freytag, Sebastian Günther, Natascha Kellner, and Julien Schuckelt for their technical assistance.

## AUTHORS CONTRIBUTION

Conceptualization CT, GRF, CG; Methodology CT, UZ, MM, CG; Experiments CT, MM; Analysis CT; Writing original draft CT, CG; Review and Editing all authors.

## COMPETING INTERESTS

The authors declare no competing interests.

## Notes

### Competing Interest Statement

The authors have declared no competing interest.

### Author Declarations

All aspects were ethically approved by the ethics committee of the Medical faculty of the University Cologne (file no. 17-244) and carried out in accordance with the Declaration of Helsinki (October 2013)

