## Supplementary material for "Local Neuronal Sleep after Stroke: The role of cortical bistability in brain reorganization": Supplementary_Calimero_Stroke_Source_BrainStim_revision_final_clean.pdf

### I. Subjects

Forty patients (36 right-handed according to Edinburgh Handedness Inventory (EHI), 6 females) with unilateral upper limb motor deficit ranging from mild to severe ((Action Research Arm Test (ARAT): 18.5±22.8; 0-55; National Institutes of Health Stroke Scale (NIHSS): 7.6±3.5; 1-14) due to acute first-ever ischemic stroke were recruited from the Department of Neurology at the University Hospital of Cologne. Patients were included based on the following inclusion criteria[1]: (i) ischemic stroke as verified by diffusion-weighted magnetic resonance imaging (DWI), (iii) structurally intact ipsilesional precentral gyrus, (iv) ≤14 days elapsed from symptom onset (6.6±2.7; 2-14 days). Exclusion criteria were: (i) any contraindication to TMS, (ii) bihemispheric infarcts, and (iii) hemorrhagic stroke.

Thirty-five patients could be re-assessed clinically, twenty-eight patients participated in the TMS-EEG follow-up visit at least 3 months later (138.3±26.6; 99-183 days). Three of these patients were only assessed at the behavioral level as they could not leave their care facility. Four other patients did not agree to a second TMS-EEG session, resulting in twenty-eight patients who participated in the second TMS-EEG experiments more than three months after stroke. Fifteen age-matched healthy participants without any history of neurological or psychiatric disease (2 females, 14 right-handed) served as a control group.

### II. TMS-EEG recordings

TMS-EEG was recorded using a TMS-compatible 64-channel EEG system (BrainAmp DC, BrainProducts GmbH, Gilching, Germany). Scalp EEG was recorded by 62 TMS-compatible Ag/AgCl sintered ring electrodes mounted on an elastic electrode cap (EasyCap-Fast'n Easy 64Ch) following the standard layout and the international 10-20 system arrangement. The two remaining electrodes of the 64-

channel system were used to record horizontal and vertical eye movements. To prevent EEG auditory evoked potentials induced by the TMS click, participants wore inserted earplugs[2–4]. Despite the advantages of masking the TMS click with white noise to reduce confounds of peripheral co-activation[5], we did not employ this procedure given the high masking intensities of up to 90 dB, which proved to be very difficult to bear for acute stroke patients[4]. Importantly, using the same approach, we previously provided evidence to capture a biologically relevant neural signal and render major confounds of peripheral co-activation very unlikely[1].

During the TMS-EEG recordings, subjects were seated in a comfortable chair and were asked to stay awake with their eyes open. Severely affected patients, unable to sit in a chair due to trunk weakness, were assessed in their beds with a 45° incline of the head section, awake and with their eyes open (n=23).

### III. TMS-EEG analysis

*LMFP*

To characterize TMS-evoked EEG potentials of the ipsilesional M1, we calculated the Local Mean Field Power (LMFP) quantifying the evoked electric field as a function of time for the channels closest to the site of stimulation. The LMFP is computed as the square root of squared TEPs averaged across channels closest to the site of stimulation (FC3-FC1-C3-C1/FC4-FC2-C4-C2 for ipsilesional M1)[6,7].

*Slow wave amplitude*

To explicitly address the phenomenon of sleep-like slow waves, we further assessed the amplitude of TMS-evoked low-frequency components[8,9]. For this purpose, single trials were low-pass-filtered <4Hz, Chebyshev 3$^{rd}$ order), averaged, and rectified. For each channel, the maximum slow wave amplitude was computed within the time window of 8-350 ms post-stimulus and subsequently measures calculated at the single channel level were averaged.

*Natural frequency*

Spectral features were evaluated by computing the event-related spectral perturbation (ERSP) after time-frequency decomposition using the Morlet wavelet transform (3.5 cycles). This procedure was implemented using the EEGLAB function *newtimef*[10]. Absolute spectra normalization was applied at the single trial level, first performing a full epoch length single trial normalization[11] and subsequently by pre-stimulus baseline correction (-500--100ms) on the resulting ERSP averaged across all trials. Finally, only significant ERSP values surviving bootstrap-based statistics (number of permutations=1000, alpha<0.05) concerning the baseline were considered for analysis. We averaged the ERSP in the 5-50 Hz frequency range and in a time window between 20 and 200ms post-stimulus to minimize the effects of possible artifacts occurring at the time of stimulation[12]. For subsequent analyses, we extracted the frequency with the maximum power, i.e., the natural frequency[12], of the TMS-evoked brain response for the ipsilesional motor cortex. Besides, with explicit regard to cortical off-periods associated with slow waves, the suppression of high-frequency EEG power (>20Hz) was assessed[8,9]. Accordingly, we extracted the integral of high-frequency power between 100 and 350ms after TMS onset for the ipsilesional motor cortex[9,13].

*Phase locking factor*

Local causal interactions induced by the TMS pulse over ipsilesional M1 were quantified by means of the phase locking factor (PLF)[8,9,14], calculated for every single electrode as the absolute value of the average of the Hilbert Transform across trials. The PLF is assumed to be a proxy of connectivity evaluating the instantaneous phase difference under the hypothesis that functionally connected areas should be synchronized in response to a given input across trials in a specific time window. In this context, the time course of PLF was used to assess the duration of deterministic effects of TMS perturbation in a specific time window[9]. Here, single trials were high-pass filtered (>8Hz, Butterworth 3$^{rd}$ order) before PLF calculation. Statistical differences from baseline (−500 - −100ms) were assessed for each contact by bootstrap statistics (number of permutations=1000, alpha<0.05). PLF values not significantly different from baseline were set to zero. For each channel, the latest significant PLF time point was identified and averaged over the four channels closest to the site of stimulation (FC3-FC1-C3-C1/FC4-FC2-C4-C2 for ipsilesional M1).

*Perturbational complexity index*

To assess the complexity of global causal interactions, we calculated the perturbational complexity index (PCI), which is defined as the normalized Lempel-Ziv complexity of the spatiotemporal pattern of cortical activation triggered by a direct TMS perturbation. The PCI captures the deterministic patterns of neural activation in response to a direct perturbation, i.e., the TMS pulse[15], and directly reflects the joint ability of integration and differentiation[15,16]. Specifically, preprocessed data epoched ±400ms around the TMS onset were utilized for source modeling (three spheres BERG method as conductive head model, weighted minimum norm constraint as inverse solution), and subsequently, non-parametric bootstrap statistics were performed to extract the significant spatiotemporal patterns of TMS-evoked cortical source activity[9,15,16]. PCI was obtained by compressing the binary matrix of significant sources with the algorithmic complexity measure of Lempel-Ziv complexity and normalized by the correspondent source entropy, resulting in a positive number between 0 and 1 with PCI=1 for maximally complex TMS-evoked potentials.

### IV. Lesion mapping

For all stroke patients, magnetic resonance images (MRI) using standard sequences (DWI, T2- weighted, T2*-weighted, FLAIR, and time-of-flight angiography sequence) were acquired in a routine clinical setting within the first two days after stroke (1.5T MRI scanner; Philips, Guildford, Great Britain). Based on the diffusion-weighted images (DWI) revealing the acute ischemic lesion (TR=3900ms, TE=95ms, FOV=230mm, 22 axial slices, voxel size=1.8x2.99x6mm$^3$), individual lesion maps were created using the software MRIcron (https://www.nitrc.org/projects/mricron). After interactive delineation of individual lesion volumes, lesion maps and DWI images were co-registered to the individual FLAIR/T2-weighted images and normalized to the T2-weighted MNI template implemented in Statistical Parametric

Mapping (SPM12, http://www.fil.icon.ucl.ac.uk/). Lesions located in the right hemisphere were flipped along the mid-sagittal plane before spatial normalization.

Next, we calculated for each lesion the lesion volume, the probabilistic volume of the lesion overlap with the cortical spinal tract (CST). Moreover, we quantified the lesion effects on the brain's connectome using the Lesion Quantification Toolkit (https://wustl.box.com/v/LesionQuantificationToolkit)[17]. Accordingly, via an atlas-based approach, the parcel-level grey matter lesion load, the white matter tract disconnections, and the parcel disconnection were estimated. We, here, used the parcellation atlas consisting of 100 cortical parcels[18] plus 17 subcortical parcels from the AAL atlas[19]. Differences for these parameters between the patient subgroups, i.e., severely affected patients or patients featuring a slow wave response, were calculated by a one-way ANOVA, Bonferroni-corrected for multiple comparisons.

### V. Lesion characteristics

Estimating the parcel-level grey matter lesion load based on the 100 parcellation Schaefer atlas[18] yielded no difference in lesion-induced damage of anatomically or functionally defined brain regions for patients with or without slow wave responses (all p-values>0.2). Likewise, lesion volume was not significantly different (all p-values>0.2). This finding implies that slow waves and cortical off-periods after stroke are not merely a consequence of lesion load. Of note, while severely affected patients showed increased disconnection severity for the CST ($p<0.001$, $t_{(39)}=3.66$), the patients with sleep-like waves did not ($p=0.2$). Correspondingly, the weighted probabilistic CST overlap was solely higher in severely affected patients ($p=0.002$, $t_{(38.7)}=3.46$) but not in the slow wave group ($p=0.15$).

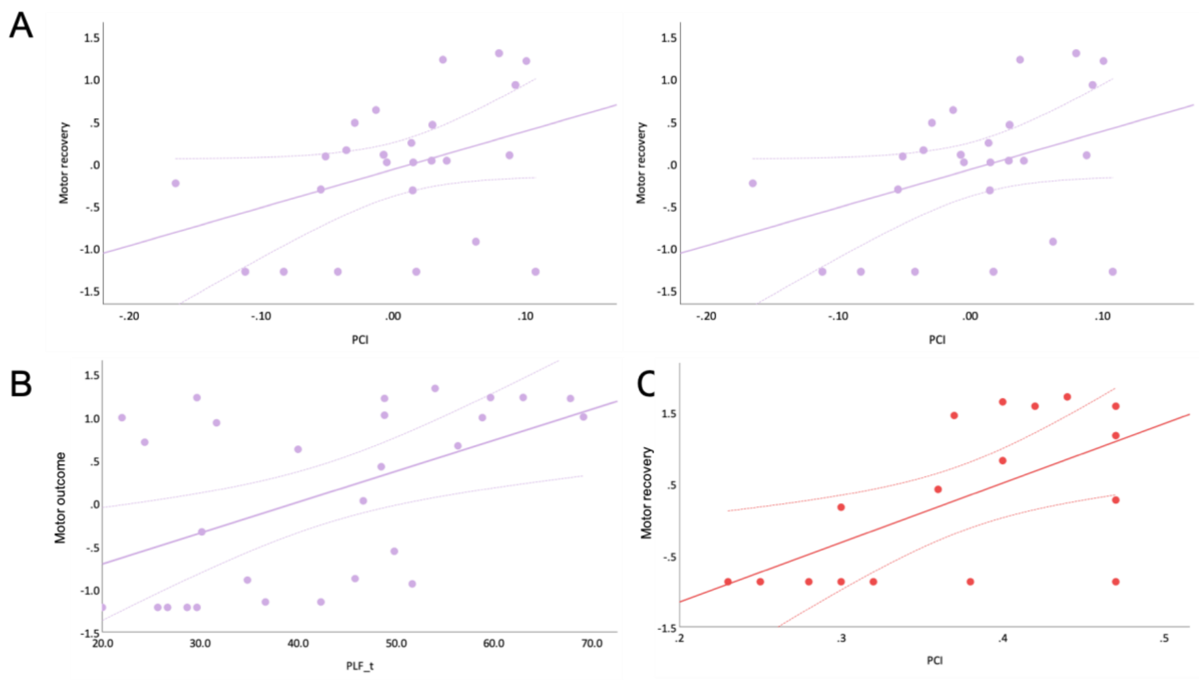


Supplementary Figure 1 – Associations between TMS-EEG parameters and motor outcome

(A) Stepwise linear regression analysis revealed the initial motor deficit, as indexed by the motor composite score, and PCI as the predictors to explain motor recovery in the chronic phase after stroke ($R^2$=0.49, adjusted $R^2$=0.44, $F_{(2,22)}$=9.50, p=0.002). Please note that the scatterplots are showing the standardized residuals of the regression's predictors. (B) Persistent alterations of the causal effects on TMS-evoked local cortical interactions ($PLF_t$) in the second session post-stroke was related to the residual motor deficit (r=0.59, p<0.001). (C) For the subgroup of MEP negative and severely affected patients, PCI was the sole predictor of motor recovery ($R^2$=0.65, adjusted $R^2$=0.60, $F_{(1,9)}$=12.01, p=0.016). Please also note that motor outcome and motor recovery are shown as the PCA-generated composite score. Lines indicate linear regression lines and curves the 95% confidence interval.
